# The association between rs6859 in *NECTIN2* gene and Alzheimer’s disease is partly mediated by pTau

**DOI:** 10.1101/2024.06.21.24309310

**Authors:** Aravind Lathika Rajendrakumar, Konstantin G. Arbeev, Olivia Bagley, Anatoliy I. Yashin, Svetlana Ukraintseva

## Abstract

**Introduction:** Emerging evidence suggests a connection between vulnerability to infections and Alzheimer’s disease (AD). The nectin cell adhesion molecule 2 *(NECTIN2)* gene coding for a membrane component of adherens junctions is involved in response to infection, and its single nucleotide polymorphism (SNP) rs6859 was significantly associated with AD risk in several human cohorts. It is unclear, however, how exactly rs6859 influences the development of AD pathology. The aggregation of hyperphosphorylated tau protein (pTau) is a key pathological feature of neurodegeneration in AD, which may be induced by infections, among other factors, and potentially influenced by genes involved in both AD and vulnerability to infections, such as *NECTIN2*.

**Materials and methods:** We conducted a causal mediation analysis (CMA) on a sample of 708 participants in the Alzheimer’s Disease Neuroimaging Initiative (ADNI). The relationship between rs6859 and Alzheimer’s disease (AD), with AD (yes/no) as the outcome and pTau-181 levels in the cerebrospinal fluid (CSF) acting as a mediator in this association, was assessed. Adjusted estimates from the probit and linear regression models were used in the CMA model, where an additive model considered an increase in dosage of the rs6859 A allele (AD risk factor).

**Results:** The increase in dose of allele A of the SNP rs6859 resulted in about 0.144 increase per standard deviation (SD) of pTau-181 (95% CI: 0.041, 0.248, p<0.01). When included together in the probit model, the change in A allele dose and each standard deviation change in pTau-181 predicted 6.84% and 9.79% higher probabilities for AD, respectively. In the CMA, the proportion of the average mediated effect was 17.05% and was higher for the risk allele homozygotes (AA), at 19.40% (95% CI: 6.20%, 43.00%, p<0.01). The sensitivity analysis confirmed the evidence of a robust mediation effect.

**Conclusion:** This study reported a new causal relationship between pTau-181 and AD. We found that the association between rs6859 in the *NECTIN2* gene and AD is partly mediated by pTau-181 levels in CSF. The rest of this association may be mediated by other factors. Further research, using other biomarkers, is needed to uncover the remaining mechanisms of the association between the *NECTIN2* gene and AD.

## 1 Introduction

Alzheimer’s disease (AD) is a debilitating neurodegenerative disorder that causes severe cognitive impairment **(Filho et al., 2017)**. Identification of biomarkers that accurately reflect pathological changes in the brain is critical for AD prediction and treatment **(Cox et al., 2022)**. Protein modification known as hyperphosphorylation of Tau proteins (pTau) contributes to neurofibrillary tangles, a central feature of Alzheimer’s pathology **(Drummond et al., 2020)**. The pTau concentrations were also linked to amyloid-β, another main AD biomarker, in a dose-response manner **(Hirota et al., 2022)**. The utility of pTau as a sensitive marker of AD has been well demonstrated **(Moore, Hung, and Fortin 2023)**. Some studies also suggested that the aggregation of pTau may be induced by infections, among other factors **(Lee et al., 2022; Sathler et al., 2022; Shen et al., 2022; Tang et al., 2022)**. The involvement of infections in AD is an emerging field of AD research, and the literature on the factors that may influence both vulnerability to infections and neurodegeneration is increasing **(Yashin et al., 2018; Yong et al., 2021; Cairns, Itzhaki and Kaplan, 2022; Mancuso et al., 2022; Nuovo et al., 2022; Piekut et al., 2022; Sathler et al., 2022; Tang et al., 2022; Goldhardt et al., 2023; Ukraintseva et al., 2023; Popov et al., 2024)**. Nectin-2 (Nectin cell adhesion molecule 2) is a membrane component of adherens junctions that serves as an entry point for certain herpesviruses and may mediate response to the viral infection **(Ogawa et al., 2022; Goldhardt et al., 2023)**. The single nucleotide polymorphism (SNP) rs6859 of the *NECTIN2* gene was highly significantly associated with AD risk in observational human data **(Yashin et al., 2018; Mizutani et al., 2022)**. However, it is unclear how exactly rs6859 facilitates the development of AD pathology. It is unlikely to be through the effects of *APOE4* (located nearby, on the same chromosome 19), because rs6859 is not in the linkage disequilibrium with *APOE4*. It is possible that the *NECTIN2* polymorphism contributes to AD through its involvement in vulnerability to infections. Since pTau levels can be influenced by infections and related factors, we hypothesized that they may also be influenced by the *NECTIN2* gene, involved in both AD and vulnerability to infections, and could mediate the association of this gene with AD.

In this paper, we performed causal mediation analysis (CMA) to explore causal relationships among the rs6859 polymorphism in the *NECTIN2* gene, pTau, and AD using the Alzheimer’s Disease Neuroimaging Initiative (ADNI) data.

## 2 Materials and methods

### 2.1 Data

We used the UPENNBIOMK Master dataset publicly available from the ADNI consortium upon request (<u>adni.loni.usc.edu</u>). ADNI is an ongoing research study envisaged under Michael W. Weiner with the original aim of generating data from multiple biomarkers, along with clinical and other cognitive assessments, to learn about mild cognitive impairment (MCI) and its conversion to full-blown AD in US older adults. The minimum age of recruitment was 55 years, and the study tracks individuals with AD, cognitively normal, and milder cognitive deficits across time **(Weiner et al., 2017)**. The database currently contains comprehensive measurements of various biomarkers, genetic variation, brain volume and structure, and cognitive status (**Weiner et al., 2010**). The ADNI was conducted in 3 waves: ADNI 1, ADNI GO, ADNI 2, and ADNI 3. For these phases, different genotyping platforms were used for genome-wide association studies (GWAS). These were Illumina Human610-Quad BeadChip, Illumina HumanOmniExpress BeadChip, and Illumina Infinium Global Screening Array v2 (GSA2), respectively, for ADNI waves 1-3. Biomarkers and cognitive measurements were collected during the baseline and subsequent visits. The recorded number of visits ranged from a single visit to a maximum of seven visits per participant. The ADNI data contain individual measurements of pTau-181 levels in the cerebrospinal fluid (CSF) extracted using lumbar puncture and quantified by the fully automated mid-region Roche Elecsys electrochemiluminescence immunoassays platform. This pTau181 measure is considered a reference assay for comparing the performance of new pTau epitopes in differentiating AD progression **(Suárez Calvet *et al*., 2020)**. We extracted the pTau-181 information with the corresponding subject ID (RID) and sample draw date and linked the files with age and other demographical information from the demography file. The genotypes of SNP rs6859 were extracted from the Plink binary files using the --recode command from the Plink 1.90 beta version **(Purcell and Chang, 2023.; Chang et al., 2015)**. The SNP information file was used to cross-check the genotype coding. We used the ADNIMERGE file to determine the case and control status for AD.

### 2.2 The Causal Mediation Analysis (CMA)

In this analysis, we hypothesized that the effect of the *NECTIN2* polymorphism, represented by the SNP rs6859, on AD is mediated through the pTau levels measured in the CSF. The CMA is a common method for dissecting the total effect of treatment into direct and indirect effects **(Nguyen, Schmid and Stuart, 2020).** The indirect effect is transmitted via a mediator to the outcome (Figure illustrating the CMA approach in the supplement). The CMA will allow us to see if, and to what degree, the rs6859 could indirectly facilitate AD occurrence through the pTau. In other words, what part of the association between rs6859 and AD could be explained by the causal connection between pTau and AD.

### Assessment of rs6859, pTau-181, and AD relationship

In this CMA, the relationship between rs6859 and AD, with AD (yes/no) as the outcome, and pTau-181 levels (pg/mL) in the cerebrospinal fluid (CSF) acting as a mediator in this association, was assessed. The pTau-181 was a reasonable choice as a mediator variable for our CMA also because it allowed discrimination between pathology-confirmed AD and other dementia subtypes **(Grothe et al., 2021)**. Information on AD status (yes/no) was an outcome. The control group included both cognitively normal individuals and subjects with dementia, but not AD.

### Statistical Analysis

We used R software version 4.0.0 (Vienna, Austria) to perform the analysis **(R Core Team 2021)**. *ggplot2* and *MuMin* packages were used for creating plots and regression model selection, respectively **(Wickham, 2016; Bartoń, 2013)**. Continuous variables with a normal distribution were summarized as mean ± standard deviation (SD) or median and interquartile range (IQR) for visually skewed distributions. For the causal mediation effects analysis, an additive effect of the increasing dose of allele A of the SNP rs6859 was considered in the model. For participants, different concentrations of pTau-181 were recorded during the same visit. To quantify the variation for pTau-181, we calculated the overall median CV% change for the sample and the coefficient of variation percentage (CV%) for repeat measures from the same day, along with their bootstrapped confidence intervals (CI). For the corresponding measures, CV% was calculated by dividing the standard deviation by the mean and then multiplying by 100. We did not employ a pass/fail criterion for the inclusion of values, as the variations could be biologically relevant, and pruning the data could remove important information. To understand these variations, boxplots were created for daily CV% variations across categorical variables. The pTau-181 was summarized to a single reading per participant (median value) to obtain a more stable measure and take advantage of multiple measures. pTau-181 was standardized, i.e., scaled and centred for regression and mediation analyses. Age was computed from the final pTau-181 sample draw date and the birth date.

Separate regression models were fit to assess the association between pTau-181, rs6859, and AD. We conducted a probit regression to examine the independent effect of rs6859, with the outcome adjusted for the covariates based on an Akaike Information Criterion (AIC)-informed model **(Cavanaugh and Neath, 2019)**. Alternatively, a linear regression was run similarly for estimating the pTau-181 relationship with SNP rs6859. The covariates included were age, sex, years of education, marriage status (divorced and never married vs. married), smoking (yes/no), and alcohol use (yes/no). All possible combinations of covariates were run in the model, and the model with the best AIC was selected. In the third step, the conditioned effects of rs6859 and pTau-181 on the outcome variable were estimated in a probit regression model **(Yin et al., 2016)**. Probit regression model is a non-linear model for estimating the probability of being assigned to either of the categories of a dichotomous outcome. It is similar to the logistic model except for the relationship with the link function, which is based on a cumulative Gaussian distribution **(Oyekale, 2021)**. The main advantage of the probit model over the logistic model is that the former provides the probability of the event while treating the covariate as a latent variable, while the odds ratio from the latter does not have the same interpretation **(Yin et al., 2016)**. The estimates from the probit regression model were presented as average marginal effects. Average marginal estimates represent the average conditional probability associated with the variables represented on the outcome scale **(Williams, 2012)**. Lastly, the *mediate* function from the *mediation* package was used to compute the mediation and proportion of mediated effects from the fitted models (**Tingley et al., 2014**).

We estimated the parameters by keeping the GG genotype of SNP rs6859 as the reference genotype while considering the GA/AA genotype (carriers of allele A) at greater risk for AD. These were coded as 0, 1, and 2, and the mediation effects were computed on allele (A) counts as in an additive model. We also computed the effects for AA genotype individuals while choosing the GG genotype as the reference category (GG vs. AA). The *mediation* package estimates an average effect for average causal mediation effects (ACME), average direct effects (ADE), and an average for the proportion of the mediated effects by decomposing the pre-computed SNP rs6859 and standardized pTau-181 effects on AD in the combined model estimation step. This step was required to quantify how the association between rs6859 and AD is mediated by pTau-181. ACME represents the indirect effect of the SNP rs6859 on AD routed through pTau. ADE is the effect of the SNP rs6859 on AD conditioned on the pTau-181. Similarly, the total effect is the sum of the direct and indirect effects of rs6859 on AD. The proportion of the mediated effects is a complementary parameter to the ACME, which simply quantifies the proportion of the mediated effect. A two-tailed p-value less than 0.05 was considered significant for the regression models and mediation effects. The confidence intervals were estimated with 5000 bootstrapped iterations with the *boot* option from the *mediation* package to obtain percentile confidence intervals. We conducted a sensitivity analysis using the ρ values (correlation between the residuals derived from the rs6859-AD association and rs6859 plus pTau-181, including a model for AD) plotted against ACME values with 5000 simulations using the *medsens* command from the same package to check the robustness of the results to see if they follow the sequential ignorability assumption **(Pearl 2014)**.

## 3 Results

### 3.1 Sample characteristics

All participants with complete covariate information in the database were included in the analysis. The final sample contained a total of 708 participants (Supplementary Figure S1). The pTau-181 level had a skewed distribution with a median value of 34.20 pg/mL. There was a large variation in pTau-181 between individuals (range: 6.9-213.0 pg/mL). The participant characteristics and demographic data are shown in Table 1. There were 181 AD cases, accounting for 25.56% of the total sample. The frequencies for cognitively normal individuals and individuals with cognitive deficiencies other than AD were 131 and 396, respectively. Participants were predominantly of the white race (94.06%) and males. Close to 95.90% of the participants had a marriage history, and 32.76% reported having a history of smoking. The proportion of participants with GA/AA genotypes of SNP rs6859 was 73.87%. For some participants, there were large pTau-181 variations as measured by CV% statistics (Supplementary Figures S2-S3). For the sample pTau-181 (CV%), the median was 6.08 (95% CI: 4.81, 7.15). Regarding pTau-181 (daily CV%), the median value was 1.33 (95% CI: 0.56, 2.15). When it comes to daily CV%, there was no notable difference for rs6859 polymorphisms. However, daily CV% variations were larger for males, individuals of white race, married participants, and those with a history of smoking and alcohol use (Supplementary Figure S4).

**Table 1.** Characteristics of participants in the study sample (n=708)

| Parameter | Mean (Frequency) | SD | Range |
| --- | --- | --- | --- |
| Age (Years) <sup>#</sup> | 74.19 | 7.19 | 55.33-90.79 |
| Male (%) | 399 (56.36%) |  |  |
| Female (%) | 309 (43.64%) |  |  |
| Education (Years) | 15.94 | 2.87 | 4-20 |
| **Marriage Status** |  |  |  |
| Ever | 679 (95.90%) |  |  |
| Never | 28 (3.95%) |  |  |
| Unknown | 1 (0.14%) |  |  |
| **Race** |  |  |  |
| White | 666 (94.06%) |  |  |
| Other | 42 (5.93%) |  |  |
| Smoking (Ever) | 232 (32.76%) |  |  |
| Smoking (Never) | 476 (67.24%) |  |  |
| Alcohol (Ever) | 29 (4.10%) |  |  |
| Alcohol (Never) | 679 (95.90%) |  |  |
| pTau-181 (pg/mL) <sup>#</sup> | 34.20 (95% CI: 32.0, 35.1) | 26.17 | 6.9 - 213.0 |
| pTau-181 (CV%) <sup>#</sup> | 6.08 (95% CI: 4.81, 7.15) | 17.15 | 0.0 - 77.6 |
| pTau-181 (daily CV%) <sup>#</sup> | 1.33 (95%: 0.56, 2.15) | 8.37 | 0.00 - 82.68 |
| **SNP rs6859 (Genotype Frequencies)** |  |  |  |
| GG | 185 (26.12%) |  |  |
| GA | 357 (50.42%) |  |  |
| AA | 166 (23.44%) |  |  |
| **SNP rs6859 (Allele Frequencies) <sup>\$</sup>** | | | |
| G | 51.34% |  |  |
| A | 48.66% |  |  |

Note. Data are presented as mean ± standard deviation (SD) or percentage (%) for continuous and categorical variables, respectively; <sup>#</sup> Age and pTau-181 are presented as median and IQR due to skewness of distributions. 95% CI for median was calculated using the bootstrap method. <sup>\$</sup>Allele frequencies show the frequencies of SNP rs6859 alleles in the analysed sample.

The boxplot in Figure 1A illustrates the pTau-181 distribution for males and females. Females had relatively higher median pTau-181 levels compared to males (35.1 vs. 33.0). GA carriers constituted the highest number (357, or 50.42%) among the SNP rs6859 genotypes. The pTau-181 variation by age for carriers (GA/AA) and non-carriers (GG) of SNP rs6859 allele A are shown in Figure 1B. The Pearson correlation heatmap for all continuous variables is shown in Figure 2. There were no notable, strong correlations observed among the variables. SNP rs6859 was positively correlated with pTau-181, although this correlation was relatively modest. The SNP had a weak correlation with age (0.014) and education (−0.042), respectively.

**Figure 1.**
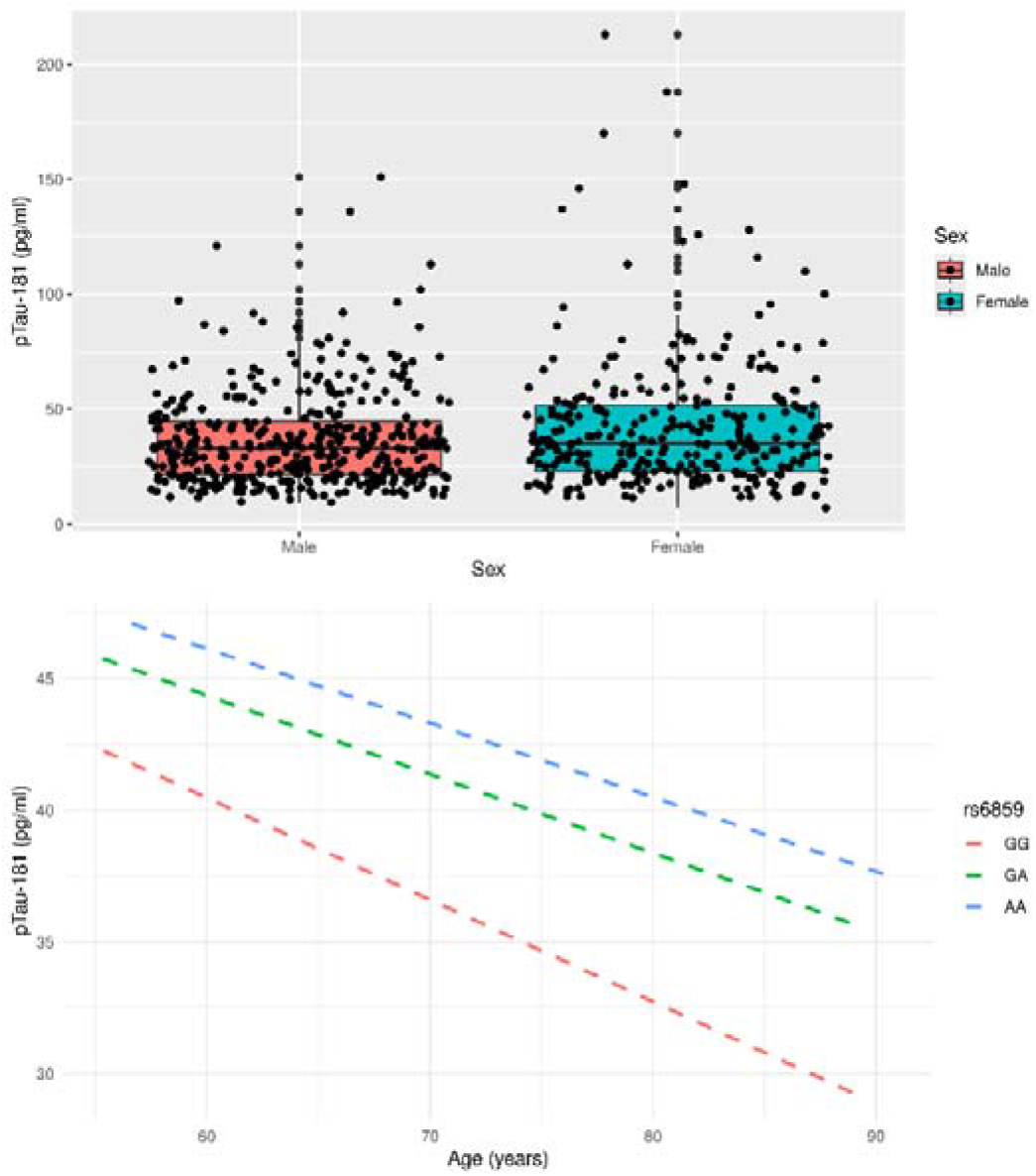
(A) Distribution of pTau-181 by gender in the ADNI cohort (B) Variation of pTau-181 distribution by age and rs6859 genotype in the study sample

**Figure 2.**
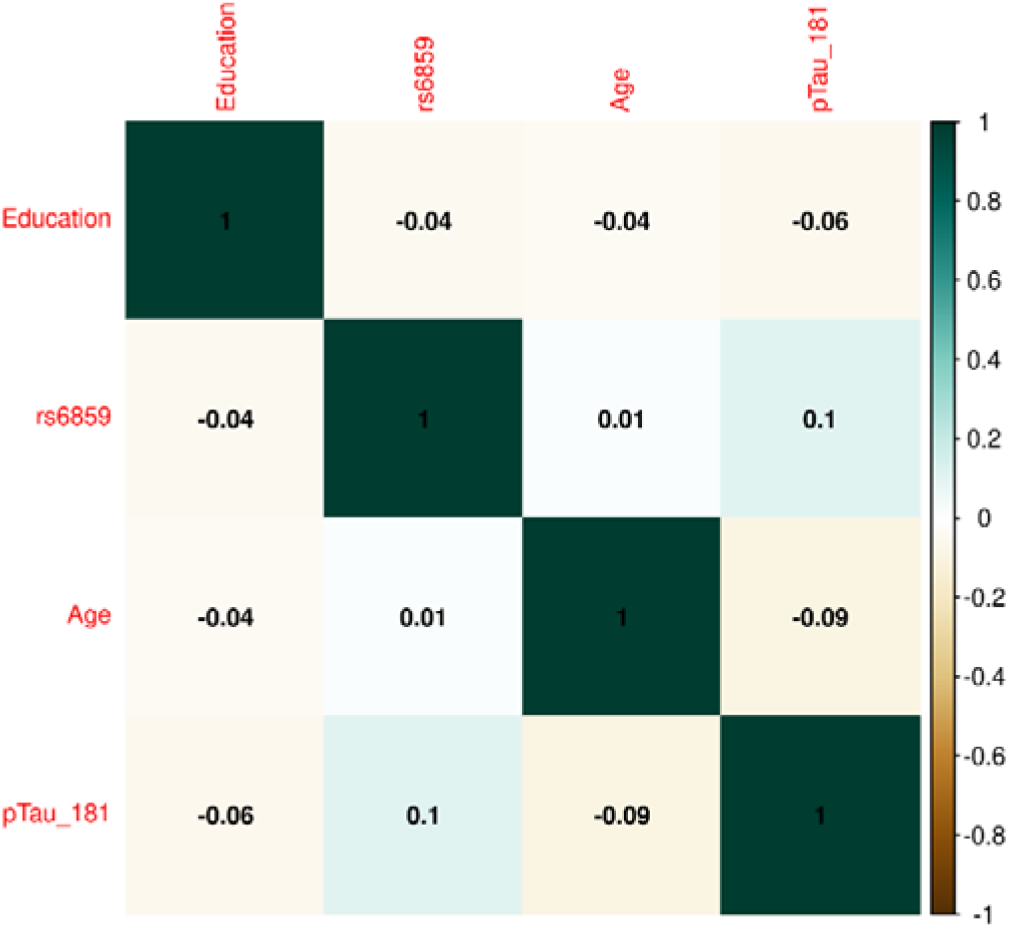
Correlation plot showing the relation for rs6859 with pTau-181 and other covariates in the ADNI dataset. Note. The size and colour of the circle indicate the strength and direction of the correlation between variables. The large dark blue dots show maximum correlation, i.e., correlation between the same variables in this case.

### 3.2 The association of the SNP rs6859 with AD

We found a statistically significant association between the SNP rs6859 and AD in the adjusted probit model. We compared the model with a complete set of covariates to the best model, which is the model with the minimal set of variables that best fits the data. The AIC - based model selection process is shown in Figure 3, which illustrates the best model selection using the Akaike weights interpreted as the conditional probabilities pertaining to the different combinations of the covariates **(Wagenmakers and Farrell 2004)**. Of these, the best model identified contained rs6859 and education (Supplementary Table S1). The AIC difference between the models was close to 10 (Supplementary Table S2). The predicted z-score from the probit regression was 0.25 (95% CI: 0.11, 0.39, p<0.001), which conveyed a higher probability of being classified as AD for the SNP rs6859 additive model (A risk allele). Thus, the first requirement of the rs6859, which is that the former should be significantly associated with both AD and pTau-181, is satisfied. The converted probit coefficients of average marginal effects for the full model are presented in Supplementary Figure S5. The average marginal effect estimates the percentage probability from the probit coefficients while keeping the effects from other variables constant. As per the average marginal effects from the best model, there was a 7.9% higher percentage probability of 0.079 (95% CI: 0.035,0.124, p<0.001) for AD associated with the genotype change from GG to AA in the SNP rs6859. The only other important variable in this model was increasing years of education. which showed a 1.3 decreased percentage probability of being classified as AD, −0.013 (95% CI: −0.024, −0.002, p<0.05).

**Figure 3.**
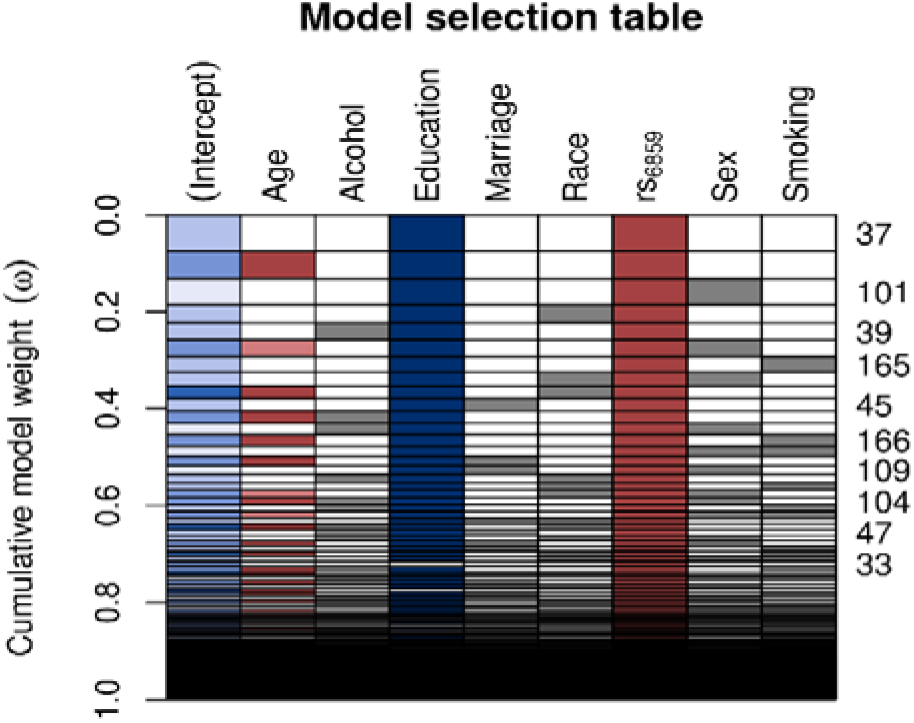
Model selection table depicting the AIC based variable selection process for the SNP rs6859 with AD. Note. The plot shows the model selection process in the automated model, ranked using AIC. Several models were run for different combinations of covariates (right-hand x-axis shown in each row), and a best subset model was selected. The models with a higher cumulative model weight indicate the best models. The best models are ranked in ascending order on the right-hand side of the plot. The row colours represent coefficient values, following a schema akin to a correlation heatmap, where blue signifies negative values and red denotes positive values. The model 37 carries the maximum weight of 0.075. The white row represents blank cells.

### 3.3 The association of SNP rs6859 with pTau-181

For evaluating the model associations for SNP rs6859 with pTau-181, we ran linear regression models. The model selection using Akaike weights is illustrated in Figure 4. SNP rs6859, age, race, and sex were selected as the most significant predictors of pTau-181 in the best subset model. The AIC difference between the models favored the model with a reduced set of covariates. The AIC for the different models is given in Supplementary Table S3. In the age and sex-adjusted model, an increase in the dose of the A allele of the SNP rs6859 was associated with an increase in the pTau-181 level. Respective coefficients and confidence limits for both models are presented in Figure 5. Our analysis of the association between rs6859 and standardized pTau-181 levels was statistically significant in the multivariate model and, therefore, independent of other risk factors, such as age and smoking. The genotype change (GG to AA) of the rs6859 resulted in about a 0.144 increase per SD of pTau-181 (95% CI: 0.041, 0.248, p<0.01). Female participants were more likely to accumulate pTau-181 than males. pTau-181 accumulation was comparatively higher in white individuals (0.277, 95% CI: −0.031, 0.586, p=0.078). However, the coefficient was not statistically significant. Surprisingly, a year of the increase in age was associated with a reduction in pTau-181 (−0.011, 95% CI: −0.021, −0.001; p<0.05). Therefore, we ran a linear mixed model with a larger sample size (records = 2189, n=1249) to confirm the association (Supplementary Figure S6). Age, gender, education, and race were adjusted in the analysis. This dataset did not include rs6859 information. Here, an increase in age predicted a 0.008 increase in pTau-181 (95% CI: 0.001, 0.015, p<0.05). Our findings show that the diminished association for pTau-181 with age likely occurred as a result of the smaller sample size arising from linking covariate information.

**Figure 4.**
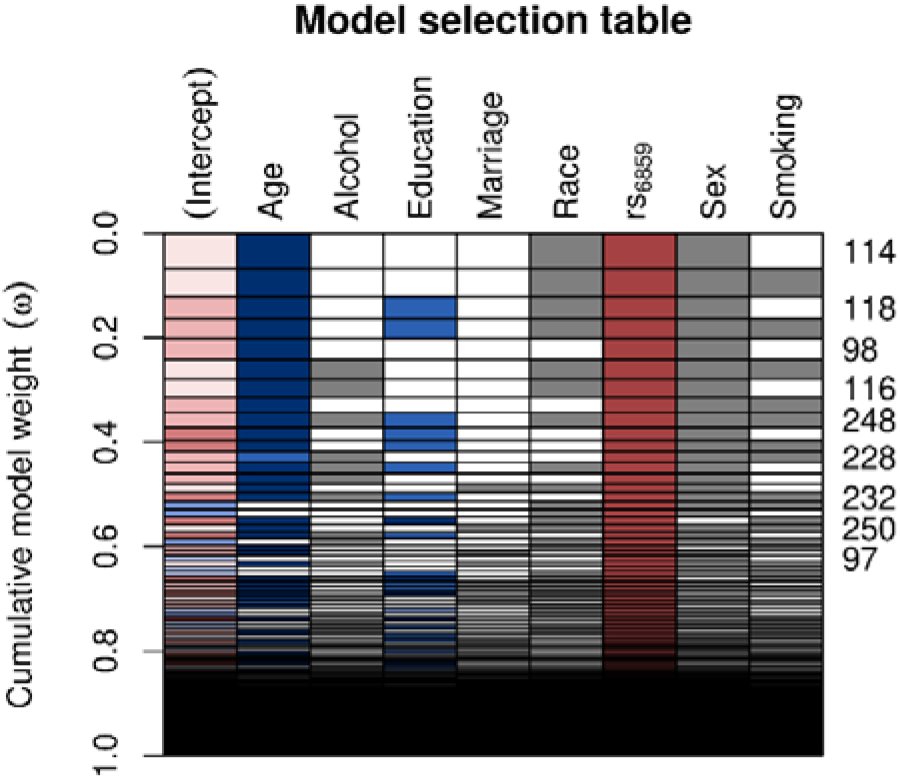
Model selection table depicting the AIC-based variable selection process for the rs6859 association with standardized pTau-181 (n=708). The plot shows the model selection process in the automated model, ranked using AIC. Several models were run for different combinations of covariates (right-hand x-axis shown in each row) and the best subset model was selected. The models with the higher cumulative model weights indicate the best model. The best models are ranked in ascending order on the right-hand side of the plot. Colors depict coefficient values, following a schema akin to a correlation heatmap, where blue signifies negative values and red denotes positive values. Grey color is used to represent categorical variables. The model 114 carries a maximum weight of 0.066. The white row represents blank cells.

**Figure 5.**
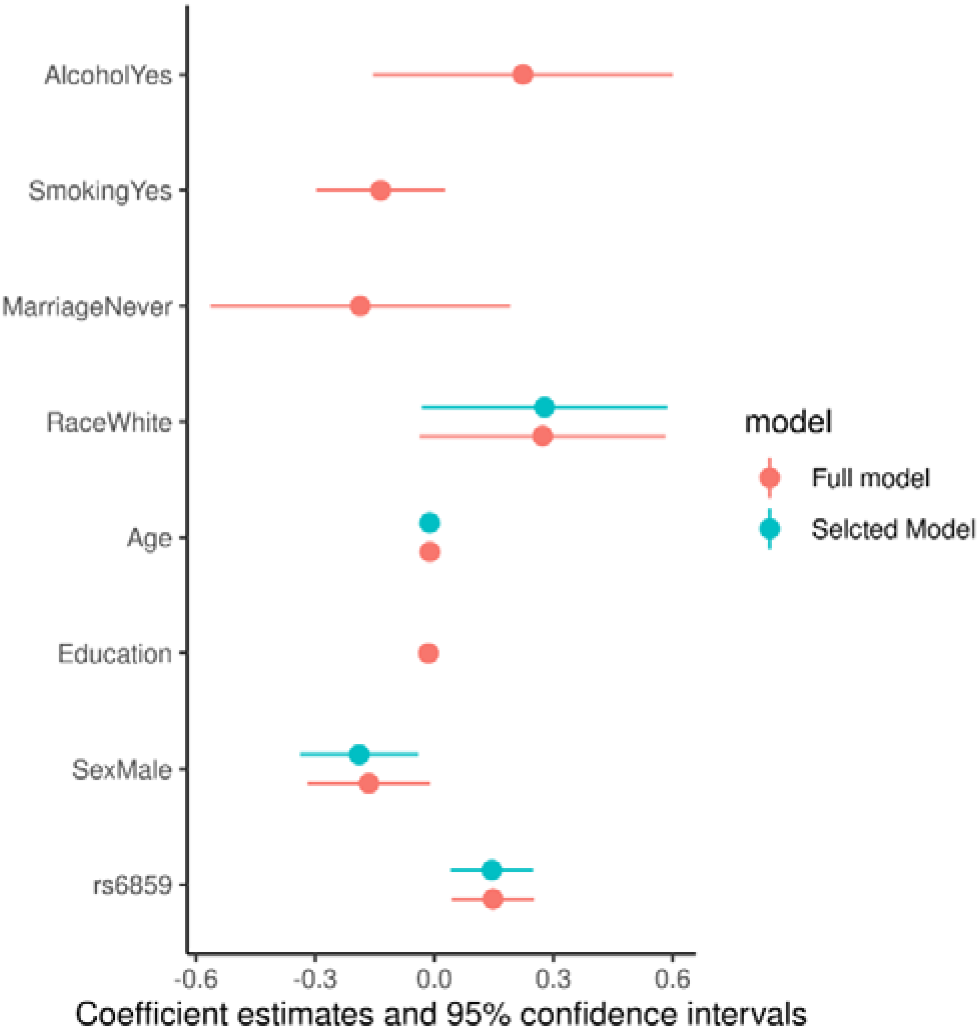
Coefficient estimates for the full and selected linear regression models for the pTau-181

### 3.4 Associations between the outcome, treatment variable, and dependent variable

For this analysis, we included the SNP rs6859 with standardized pTau-181 as covariates to generate estimates to be used later in the causal mediation analysis. The predicted z-score from the probit model was significant for both the rs6859 and mediator variables. The allele changes from G to A for the SNP rs6859 remained similar to the previously computed estimate for AD (0.229, 95% CI: 0.081, 0.378, p<0.01). For each standard deviation change in pTau-181, there was a higher z-score, implying a higher probability of being classified as AD (0.328, 95% CI: 0.225, 0.434, p<0.001). Associated probabilities for SNP rs6859 and pTau-181 were 6.84% (95% CI: 2.49, 11.18, p<0.01) and 9.79% (95% CI: 6.88%, 12.70%, p<0.001), respectively. The predicted effects from this model were visualized in the accompanying plot (Figure 6), showing an increase in predicted risk associated with the unit change in rs6859 and pTau-181. As the model estimates were significant for both rs6859 and pTau-181, the relationship is suggestive of a partial mediating effect as opposed to a full mediating effect.

**Figure 6.**
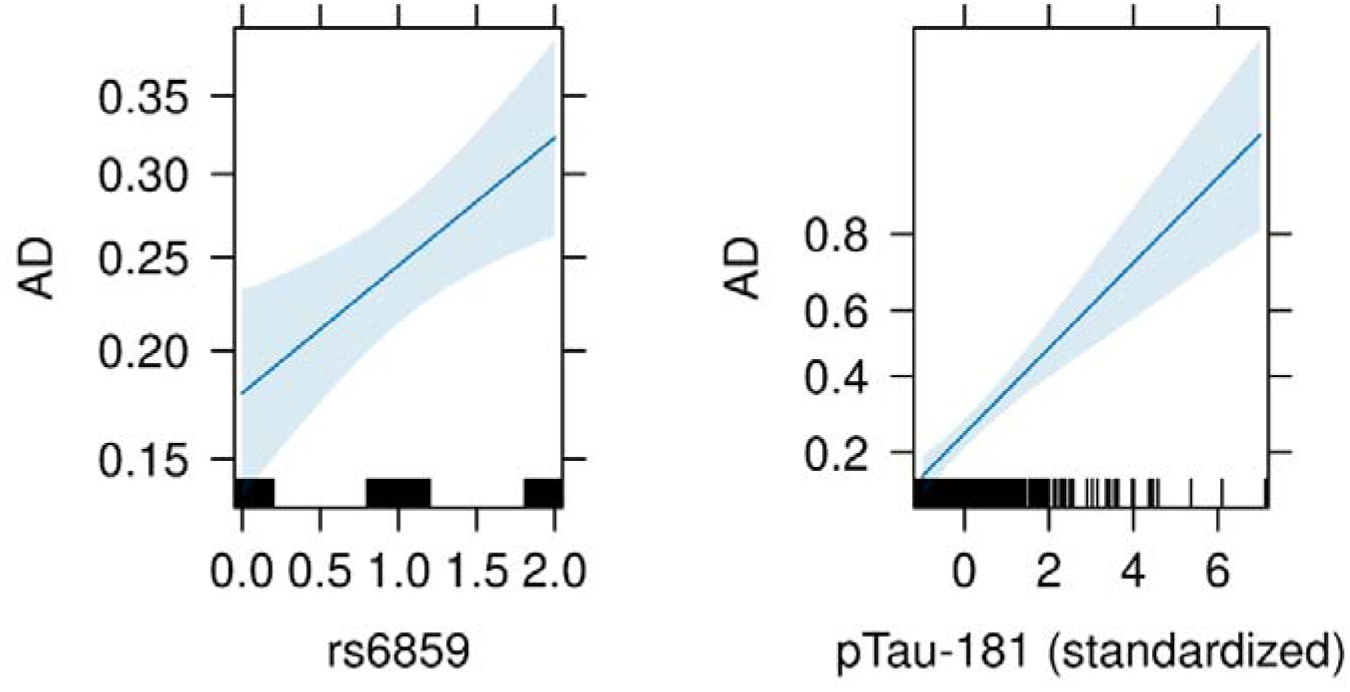
Conditioned effect plot illustrating the association of AD with rs6859 and pTau-181. Note. The plot was generated from a probit model with SNP rs6859 and pTau-181 as covariates. In Figure A, the dark blue line and surrounding light blue colour show the relationship and associated 95% confidence intervals for the genotype change and AD, respectively. In Figure B, the dark blue line shows the relationship and associated 95% confidence intervals between pTau-181 and AD. The black colour lines in the x-axis for Figure A and Figure B correspond to observed values. The y-axis shows the estimates from the probit regression. A higher positive coefficient implies a higher AD probability.

### 3.5 Findings from the causal mediation analysis

The estimands from the CMA are reported in Table 2. The variation in the SNP rs6859 predates the pTau-181 and AD. Therefore, SNP rs6859 variations could be considered a natural randomization and therefore do not have any direct influence on either the pTau-181 or AD. The estimates, as previously described, were generated from simulations, and the results indicate that the ACME, ADE, and total effect were significantly different from zero. Furthermore, the estimates could be interpreted as population averages for the specific genotypes (GG vs. AA) under a counterfactual scenario, i.e., the difference in average effects by holding the genotype effects for the AD, but switching between the values of pTau-181 observed with either of the genotypes **(Tingley et al., 2014; Zhang et al., 2016)**. As hypothesized, both the ACME and ADE were higher for the AA compared to the GG genotype and were statistically significant. The ACME for both allele variations was 0.025 and 0.033 (p<0.01), respectively. Here, ACME represents the linear chain of effects of the specific genotype on AD probability, which is indirectly affected by pTau-181. It is to be noted that pTau-181 in turn is associated with the genotype in question. The ACME estimate informs us that there is statistically significant evidence for pTau-181 mediating the observed association of SNP rs6859 with AD. On top of that, the ADE was also significant and slightly higher for the AA genotype. Therefore, this suggests that the risk predicted for allele change in SNP rs6859 was partly mediated through pTau-181. The proportion of average-mediated effect was 17.0% and was higher for the AA genotype (19.4%; 95% CI: 6.2%, 43.0%, p<0.01). For the analysis between carriers and non-carriers shown in Supplementary Figure S7, the ACME was again statistically significant and high for the carrier group (0.024 vs. 0.018, p<0.01). The corresponding proportions of the mediated effects were 18.98% and 14.56%, respectively.

**Table 2.**
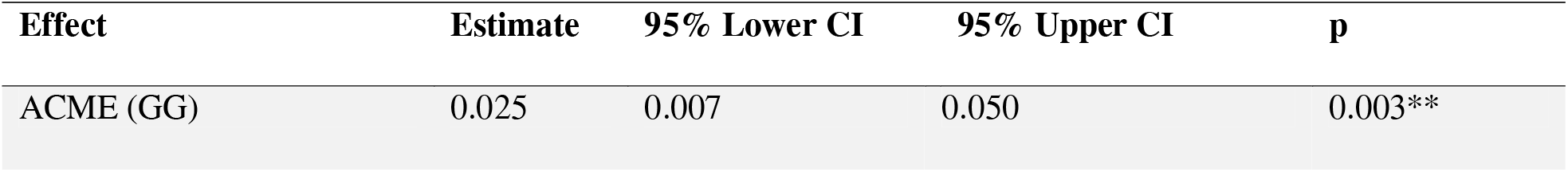
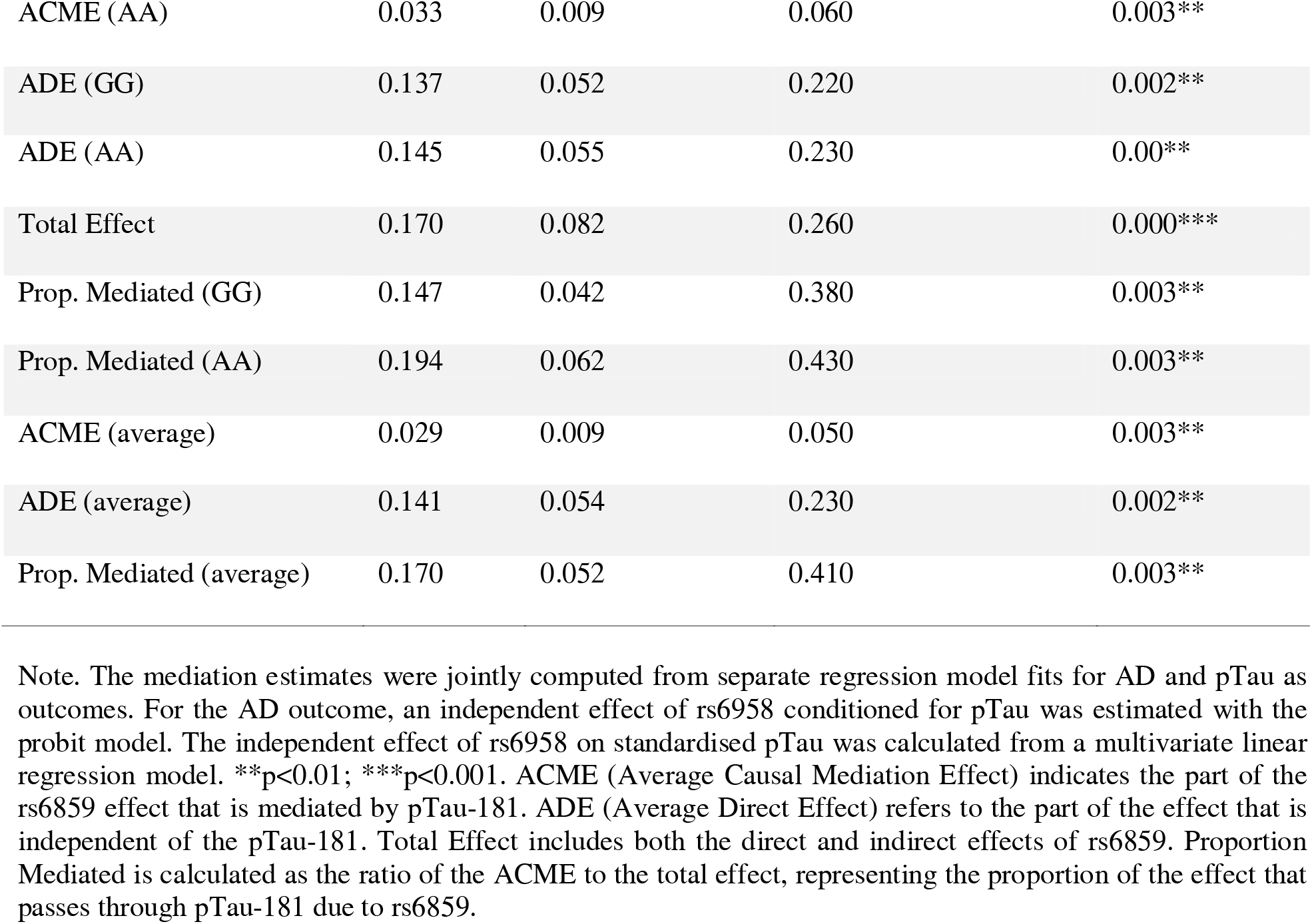
Causal mediation analysis showing the estimate of the effect on the association between rs6859 and Alzheimer’s Disease mediated through pTau-181 (n=708)

| Effect | Estimate | 95% Lower CI | 95% Upper CI | p |
| --- | --- | --- | --- | --- |
| ACME (GG) | 0.025 | 0.007 | 0.050 | 0.003** |
| ACME (AA) | 0.033 | 0.009 | 0.060 | 0.003** |
| ADE (GG) | 0.137 | 0.052 | 0.220 | 0.002** |
| ADE (AA) | 0.145 | 0.055 | 0.230 | 0.00** |
| Total Effect | 0.170 | 0.082 | 0.260 | 0.000*** |
| Prop. Mediated (GG) | 0.147 | 0.042 | 0.380 | 0.003** |
| Prop. Mediated (AA) | 0.194 | 0.062 | 0.430 | 0.003** |
| ACME (average) | 0.029 | 0.009 | 0.050 | 0.003** |
| ADE (average) | 0.141 | 0.054 | 0.230 | 0.002** |
| Prop. Mediated (average) | 0.170 | 0.052 | 0.410 | 0.003** |

Note. The mediation estimates were jointly computed from separate regression model fits for AD and pTau as outcomes. For the AD outcome, an independent effect of rs6958 conditioned for pTau was estimated with the probit model. The independent effect of rs6958 on standardised pTau was calculated from a multivariate linear regression model. \*\* $p < 0.01$ ; \*\*\* $p < 0.001$ . ACME (Average Causal Mediation Effect) indicates the part of the rs6859 effect that is mediated by pTau-181. ADE (Average Direct Effect) refers to the part of the effect that is independent of the pTau-181. Total Effect includes both the direct and indirect effects of rs6859. Proportion Mediated is calculated as the ratio of the ACME to the total effect, representing the proportion of the effect that passes through pTau-181 due to rs6859.

### 3.6 Sensitivity analysis for violation of the sequential ignorability assumption

A sensitivity analysis was carried out to check for the presence of unobserved confounders in the single mediator model (as per **Cox et al. 2013**). The results are presented in Supplementary Figure S8. For this purpose, a model correlation parameter ρ was calculated from the residuals of the models used to compute the combined model. The values of the model parameter ρ were plotted on the x-axis against ACME on the y-axis. The interpretation is based on the specific ρ values at which the ACME becomes zero or changes sign (positive to negative). The corresponding ρ values were 0.2 and 0.3 for the former and latter, respectively. According to the previous literature, the ρ values indicate that the estimands are relatively robust to the sequential ignorability assumption, and there is evidence of a positive mediation effect **(Imai and Yamamoto, 2013; Zhang et al., 2016)**.

## 4. Discussion

Herein, we investigated a possible mechanistic link between the previously observed association between the SNP rs6859 in the *NECTIN2* gene and AD, using pTau as a mediator of the rs6859 effect on AD. Tauopathies are recognised players in neurologic disorders **(Hernández and Avila, 2007)**. There is substantial evidence of associations between pTau-181 state changes and AD-relevant markers, often decades before the development of aggregated tau pathology in the brain **(Barthélemy et al., 2020)**. Despite this, it is still not clear if pTau build-up directly causes AD or if it is driven by alternative biological mechanisms **(Jack et al., 2018)**. To our knowledge, this is the first study to uncover a causal role for pTau-181 in AD from the association between variation in the *NECTIN2* gene and AD using the CMA approach. This CMA revealed that 17.05% of the rs6859 effect on AD is mediated through the pTau. Our results thus suggest that the accumulation of pTau is a *partial cause* of AD, and other, including unknown, pathways likely contribute to AD too. The latter is rather expected since AD is a heterogeneous complex health disorder and may include cases of different etiology.

Several ptau isoforms exist that could be utilized to study Alzheimer’s disease (AD) pathology **(Suárez Calvet *et al*., 2020; Hirota *et al*., 2022b; Salvadó *et al*., 2024)**. Numerous studies have explored the utility of these markers over the years, often yielding conflicting results **(Hampel *et al*., 2004; Vacchiano *et al*., 2023; Ingannato *et al*., 2024; Salvadó *et al*., 2024)**. Consequently, there is still debate regarding the best marker for AD, and new candidates are continually being proposed **(Suárez Calvet *et al*., 2020; Karikari *et al*., 2021; Salvadó *et al*., 2024)**. Assessing the diversity of ptau isoforms in relation to AD pathology is challenging, as these isoforms appear at different stages of AD and exhibit heterogeneous relationships with amyloid plaques and neurofibrillary tangles **(Wennström *et al*., 2024)**. For instance, an earlier study comparing phosphorylation at threonine 231 (p-tau231), threonine 181 (p-tau181), and serine 199 (p-tau199) in cerebrospinal fluid (CSF) showed that ptau181 could classify AD with Lewy body dementia better than p-tau231, which differentiated AD from frontotemporal dementia. However, from a clinical prediction viewpoint, these markers were only useful as standalone markers **(Hampel *et al*., 2004)**.

In mouse models, these markers were shown to be associated with synaptic dysfunction mediated by amyloid-β, with ptau181 specifically indicating axonal dysfunction (**Hirota *et al*., 2022**). Another study among Swedes demonstrated that ptau217 better reflected amyloid pathology and neurodegeneration than ptau181 and could more effectively classify AD from other neurodegenerative conditions **(Janelidze *et al*., 2020)**. Later, this finding was validated using plasma samples.**(Thijssen *et al*., 2021)**. However, both ptau181 and ptau217 share similar pathology and increase before tau accumulation in AD **(Janelidze *et al*., 2020; Horie *et al*., 2021)**. A Canadian study detected CSF-based ptau231 changes before other biomarkers in AD as a response to amyloid-β-initiated pathology **(Ashton *et al*., 2022)**. Despite their classification ability, these markers do not directly translate to sensing disease severity. For example, postmortem examinations of AD brains showed that ptau231 did not robustly indicate AD severity **(Buerger *et al*., 2002)**.

Thus, selecting the ideal ptau isoform is critical for addressing the temporality question and capturing early onset and progression in AD. We chose ptau181 as a biomarker because it is considered a “classical AD marker” and is the only ptau isoform currently measured among ADNI CSF biomarkers **(Zetterberg and Blennow, 2021)**. Plasma ptau181 levels have been demonstrated to outperform the Aβ42/Aβ40 ratio, another important AD biomarker.**(Thijssen *et al*., 2020)**. Importantly, plasma ptau181 levels can capture early dementia and are elevated in *APOE4* carriers, showing its pathological involvement in AD **(Ingannato *et al*., 2024).** Ptau181 correlates with neurodegeneration and cognitive decline in AD.**(Moscoso *et al*., 2021)**. Ptau181 assays have demonstrated reliable prediction of AD progression.**(Janelidze *et al*., 2023)**. Furthermore, CSF-based ptau181 is not influenced by other neurological conditions, unlike its plasma counterpart **(Vacchiano *et al*., 2023)**, and captures clear AD signatures compared to other dementias.**(Zetterberg, 2017)**. All this evidence shows that ptau181 is indeed a robust and tested marker of AD pathology **(Thijssen *et al*., 2021; Yu *et al*., 2023)**.

Salvadó et al. utilized a machine learning algorithm to create a biological staging model for AD using information from several biomarkers to predict AD progression. Their final set of biomarkers, however, did not include ptau181 and p-tau231, as their contribution to the model was marginal. Interestingly, the Aβ42/40 ratio and p-tau217 remained part of their list **(Salvadó *et al*., 2024)**. Therefore, more studies are required to validate and mechanistically understand how these isoforms relate to AD. Future research could use some of the markers from Salvadó et al.’s study as mediators to replicate our work. However, it is difficult to say at this stage whether these associations would yield a stronger causal relationship.

Another important gap is the lack of information regarding the tau form that can be labelled as representing potential “post-infectious dementia”. We are unaware of any specific study conducted on this topic to date. Based on current evidence, we believe that the effect of infections on AD is primarily through inflammation, which promotes AD pathological features like Aβ and ptau production **(Cairns, Itzhaki and Kaplan, 2022b; Ganz, Fainstein and Ben-Hur, 2022)**. Therefore, we hypothesize that a ptau isoform that best correlates with inflammation could be a suitable predictor.

It is important to emphasize that we selected pTau-181 as a candidate mediator of the effect of rs6859 in the *NECTIN2* gene on AD because both pTau and the *NECTIN2* gene have been linked to AD, as well as to infections, in the literature **(Yashin et al., 2018; Sathler et al., 2022; Tang et al., 2022)**. From our previous works, we also showed that upon vaccination for pneumonia and flu, rs6859 A allele carriers experienced a reduced risk for AD **(Ukraintseva et al., 2023)**. Additionally, carriers of the A allele were likely to experience cognitive decline, albeit at different rates **(Rajendrakumar et al., 2024).** In our study, the presence of the AA genotype of the SNP rs6859 was associated with an increased percentage probability for AD (by 7.9%) compared to the GG genotype, which is in line with previous findings from other cohorts **(Yashin et al., 2018; Logue et al., 2011)**. The increased pTau-181 levels in carriers of the rs6859 A allele are, however, not yet well understood. Our results do not exclude the possibility that this increase in pTau may reflect a higher vulnerability to infections as another underlying cause of AD. One hypothetical explanation could be that the variation in the *NECTIN2* gene might increase the host vulnerability to infections, e.g., herpesviruses, which in turn may promote neural damage and pTau accumulation in the brain, contributing to neurodegeneration **(Cairns, Itzhaki, and Kaplan, 2022; Powell-Doherty et al., 2020; Ogawa et al., 2022; Liu et al., 2018; Readhead et al., 2018; Goldhardt et al., 2023)**. This potential mechanism deserves further investigation.

One should note that the CMA is not the only way to uncover causal relationships from the observed associations. Mendelian randomization (MR) is another common approach. We, however, preferred the CMA for this study because it provided a more attractive option to test the putative causal pathways, given the possible role of confounding associations with the observed findings for SNP rs6859 with pTau-181 and AD **(Sait et al., 2021)**. The CMA, at least in theory, is less stringent than the assumptions associated with MR studies, which assume a much stricter instrument variable and an intermediate outcome pathway **(Rijnhart et al., 2021)**. The CMA with an ideal “treatment” should produce estimates similar to those in the MR **(Carter et al., 2021)**. Additional strengths of this study include the availability of longitudinal biomarker measurements in the ADNI and the use of sensitivity analysis. An important limitation of this study is the incomplete data on covariates for some ADNI participants, which led to a smaller sample size than initially expected. Other limitations include the lack of supporting evidence for our findings, particularly regarding how the *NECTIN2* SNP rs6859 affects the production, processing, and deposition of beta-amyloids, and its modulation of β-secretase (BACE1) activity. Furthermore, the Nectin2 protein, the end product of this gene, is not measured in ADNI. Measuring Nectin2 protein levels could provide additional downstream insights. To address these gaps, we plan to conduct future studies examining the relationship between SNP rs6859, beta-amyloids, and hippocampal volume to better understand *NECTIN2*’s impact on AD phenotypes.

In conclusion, this study revealed a new causal relationship between pTau-181 and AD that partly explains the association of the rs6859 polymorphism in the *NECTIN2* gene with Alzheimer’s disease. Our findings in ADNI data further strengthen the evidence of the association between rs6859 and AD, previously reported in GWAS of other human cohorts **(Yashin et al., 2018; Logue et al., 2011; Xiao et al., 2022)**, and provide a plausible pathway linking the *Nectin2* polymorphism with AD. Predictably, the association between rs6859 and AD was only partly mediated by pTau. The rest of the effect may be mediated through other, yet unknown, factors. Further research is needed to uncover the remaining mechanisms connecting the variation in the *NECTIN2* gene with AD.

## Data availability statement

The data used in this article is publicly accessible through the ADNI website (http://adni.loni.usc.edu).

## Ethics statement

The samples were collected with the written informed consent of all participants. Informed consent was obtained from all subjects and/or their legal guardian(s). ADNI studies follow Good Clinical Practices guidelines, the declaration of Helsinki and United States regulations (U.S. 21 CFR Part 50 and part 56). ADNI studies were approved by all the respective Institutional Review Boards of academic institutions involved in the consortium. For our specific study, we received approval from the Duke University Health System Institutional Review Board under the Protocol IDs Pro00109279 and Pro00105389.

## Author Contributions

S.U. and A.L.R. conceived the study. K.A., O.A.B., and A.L.R. were involved in data curation, analysis, and data interpretation. S.U., K.A., and A.I.Y. provided guidance on data analysis and critically revised the manuscript. All authors read and approved the final version of the manuscript.

## Abbreviations

ACME: Average Causal Mediation Effects
AD: Alzheimer’s Disease
ADE: Average Direct Effects
AIC: Akaike Information Criterion
Aβ: Amyloid βeta
BACE1: β-secretase
CV%: Coefficient of Variation Percentage
GWAS: Genome-wide association studies
HSV: Herpes Simplex Virus
IQR: Interquartile Range
pTau: Phosphorylated Tau
SD: Standard Deviation
SNP: Single Nucleotide Polymorphism
*NECTIN2*: Nectin Cell Adhesion Molecule 2 (gene)

## Acknowledgements

The authors would like to thank the ADNI team. Data collection and sharing for this project was funded by the Alzheimer’s Disease Neuroimaging Initiative (ADNI) (National Institutes of Health Grant U01 AG024904) and DOD ADNI (Department of Defense award number W81XWH-12-2-0012). ADNI is funded by the National Institute on Aging, the National Institute of Biomedical Imaging and Bioengineering, and through generous contributions from the following: AbbVie, Alzheimer’s Association; Alzheimer’s Drug Discovery Foundation; Araclon Biotech; BioClinica, Inc.; Biogen; Bristol-Myers Squibb Company; CereSpir, Inc.; Cogstate; Eisai Inc.; Elan Pharmaceuticals, Inc.; Eli Lilly and Company; EuroImmun; F. Hoffmann-La Roche Ltd and its affiliated company Genentech, Inc.; Fujirebio; GE Healthcare; IXICO Ltd.;Janssen Alzheimer Immunotherapy Research & Development, LLC.; Johnson & Johnson Pharmaceutical Research & Development LLC.; Lumosity; Lundbeck; Merck & Co., Inc.;Meso Scale Diagnostics, LLC.; NeuroRx Research; Neurotrack Technologies; Novartis Pharmaceuticals Corporation; Pfizer Inc.; Piramal Imaging; Servier; Takeda Pharmaceutical Company; and Transition Therapeutics. The Canadian Institutes of Health Research is providing funds to support ADNI clinical sites in Canada. Private sector contributions are facilitated by the Foundation for the National Institutes of Health (www.fnih.org). The grantee organization is the Northern California Institute for Research and Education, and the study is coordinated by the Alzheimer’s Therapeutic Research Institute at the University of Southern California. ADNI data are disseminated by the Laboratory for Neuro Imaging at the University of Southern California.

