## Supplementary Material for "The association between rs6859 in *NECTIN2* gene and Alzheimer’s disease is partly mediated by pTau"

**Supplementary Materials**

ADNI merge Data

(Observations=15,703, n=2375 individuals)

1654 participants were excluded:

Linked with SNP rs6859 genotype data n=930 participants, Linked with Ptau-181 data n =724

No recorded DR grade (45,5records, 66,197 PROCHI)

First screening visit >=1990-01-01 (43,6440 PROCHI, 64,879 PROCHI)

No DM diagnosis date -55728

Date out < DM diagnosis date:55616

Death date > DM diagnosis date:55612

Follow-up date > T2D date (55,327 PROCHI)

Combined data with demographic information

(n=721 individuals)

13 participants were excluded:

Linked with smoking and alcohol history data n=3,

Missing covariate data n=10

Missing covariat

Final study sample (n=708 individuals)

**Supplementary Figure S1.** Study flow diagram showing the final sample selection in the ADNI cohort

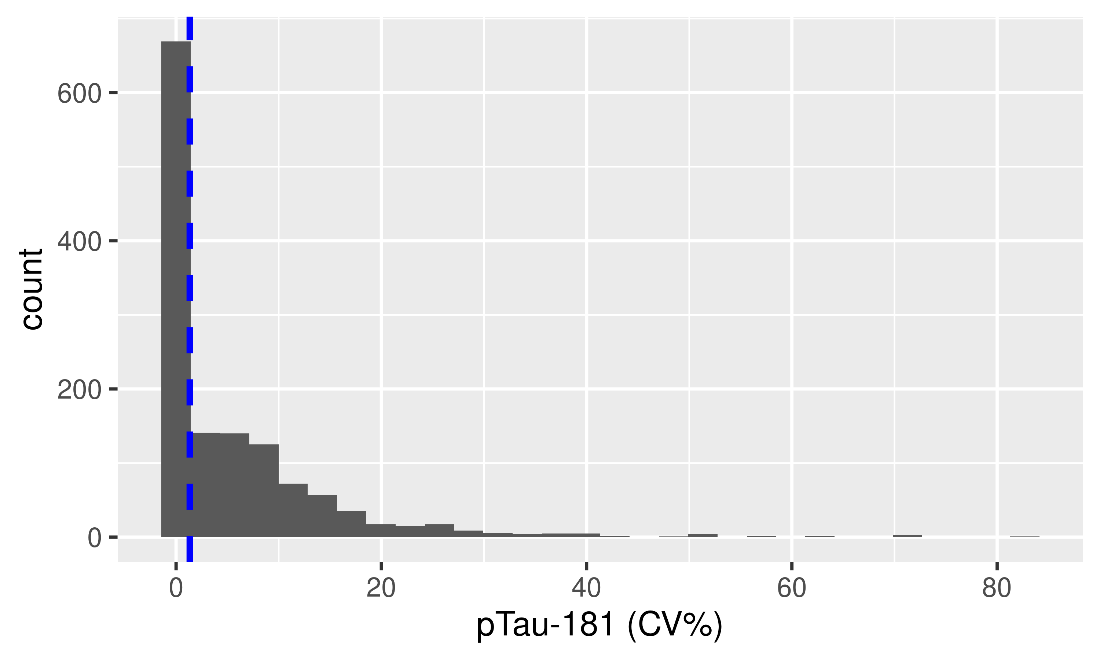

**Supplementary Figure S2.** Histogram showing daily Measurement variation for pTau-181 Indicated by CV%

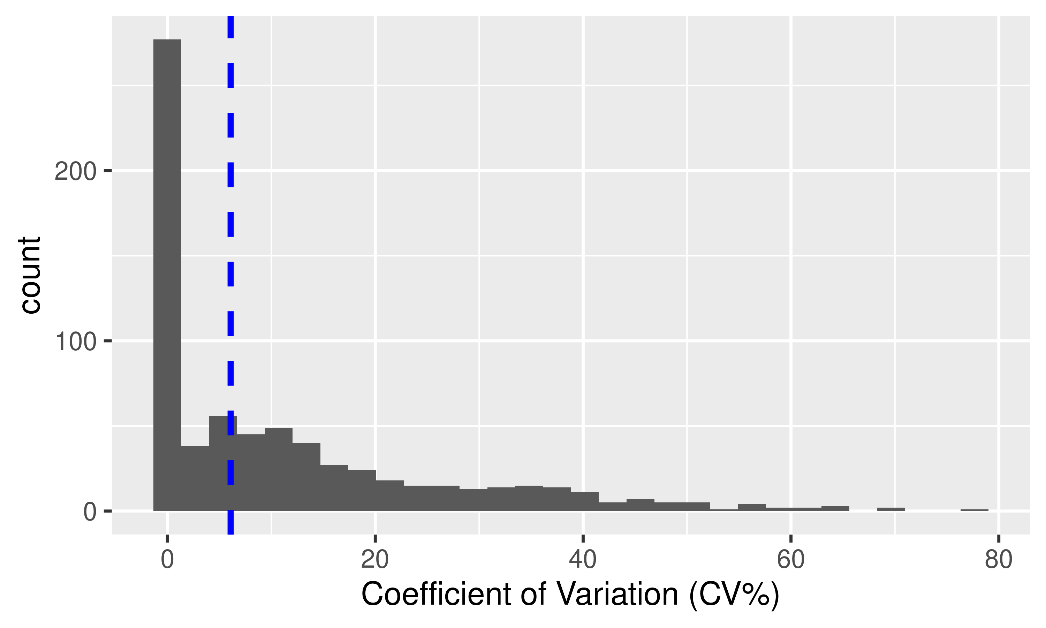

**Supplementary Figure S3.** Histogram Showing the overall sample variation for pTau-181 indicated by CV%

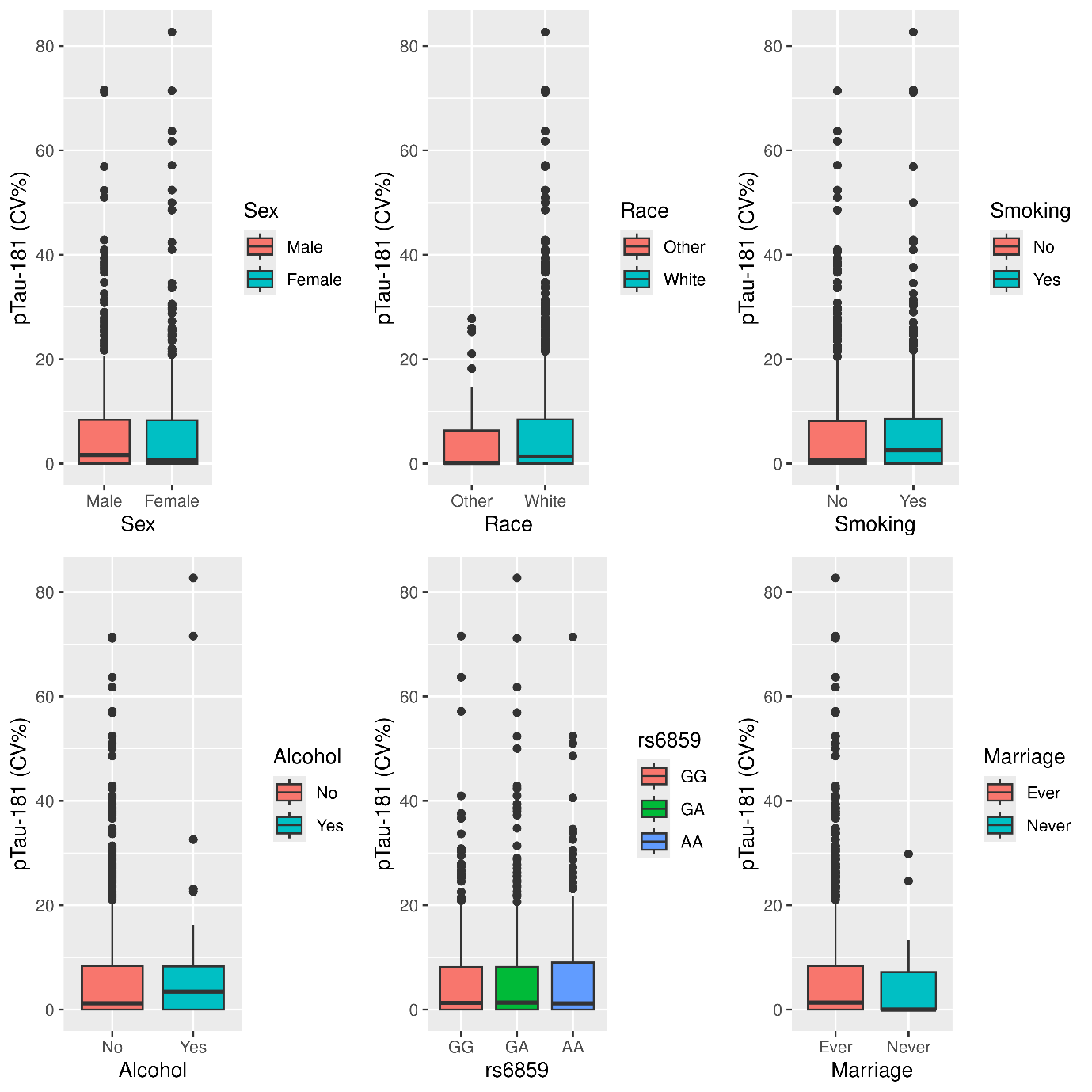

**Supplementary Figure S4.** Boxplots showing daily CV% variation of pTau-181 for categorical variables

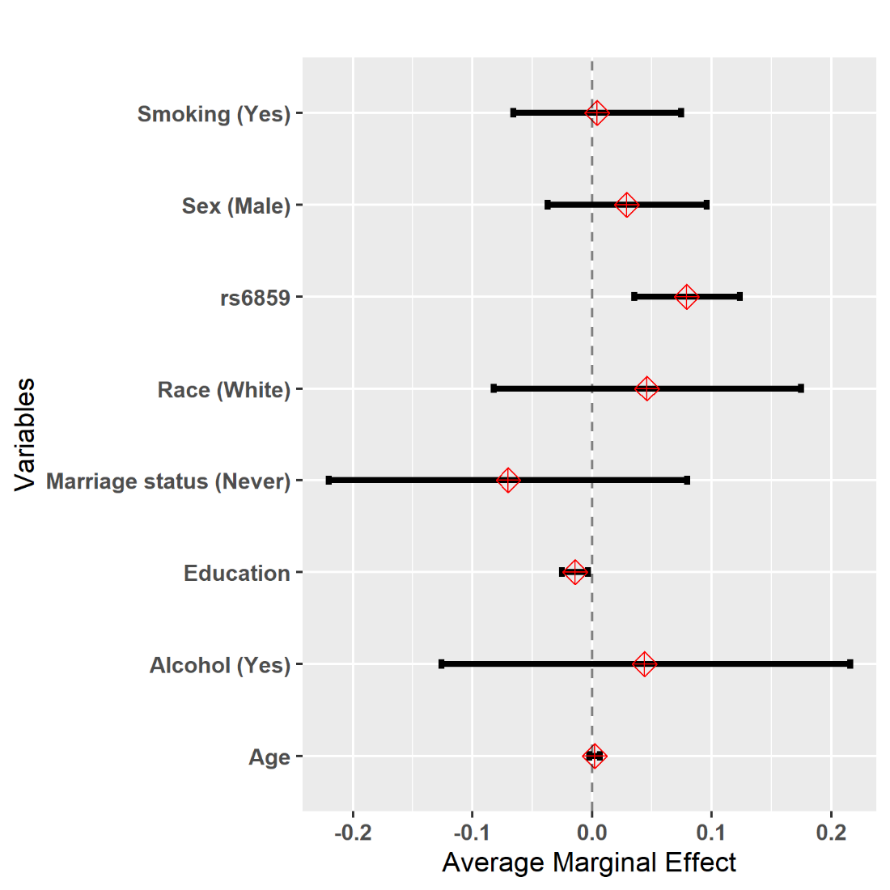

**Supplementary Figure S5.** Average marginal estimates derived from the full variable probit regression model for the prediction of AD

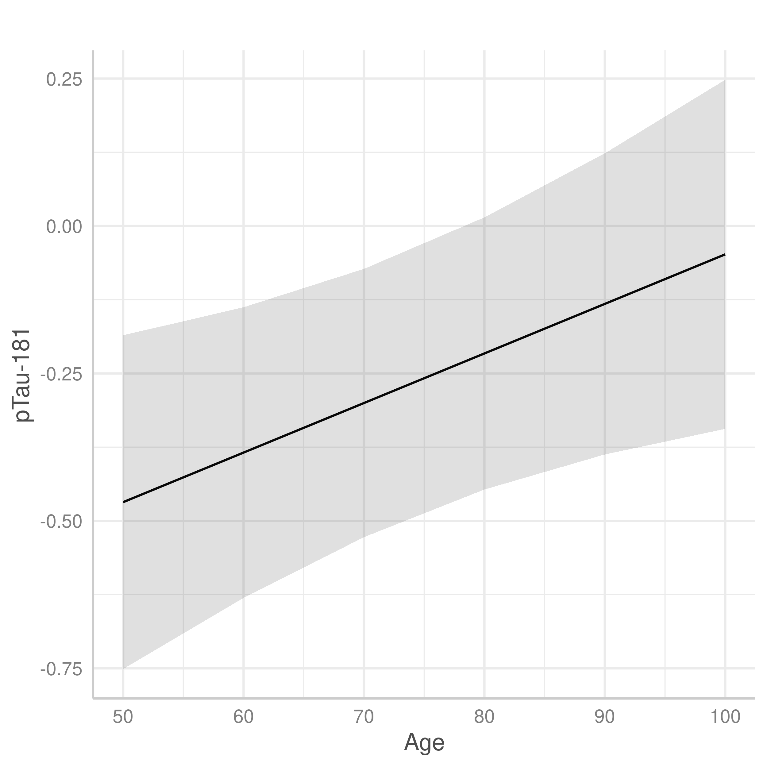

**Supplementary Figure S6.** Association of pTau-181 and age estimated from the linear mixed models with sex, race, education, and smoking history as covariates assuming varying subject-specific intercepts. Note. The solid black line and surrounding gray colour denotes the relationship and 95% confidence intervals respectively.

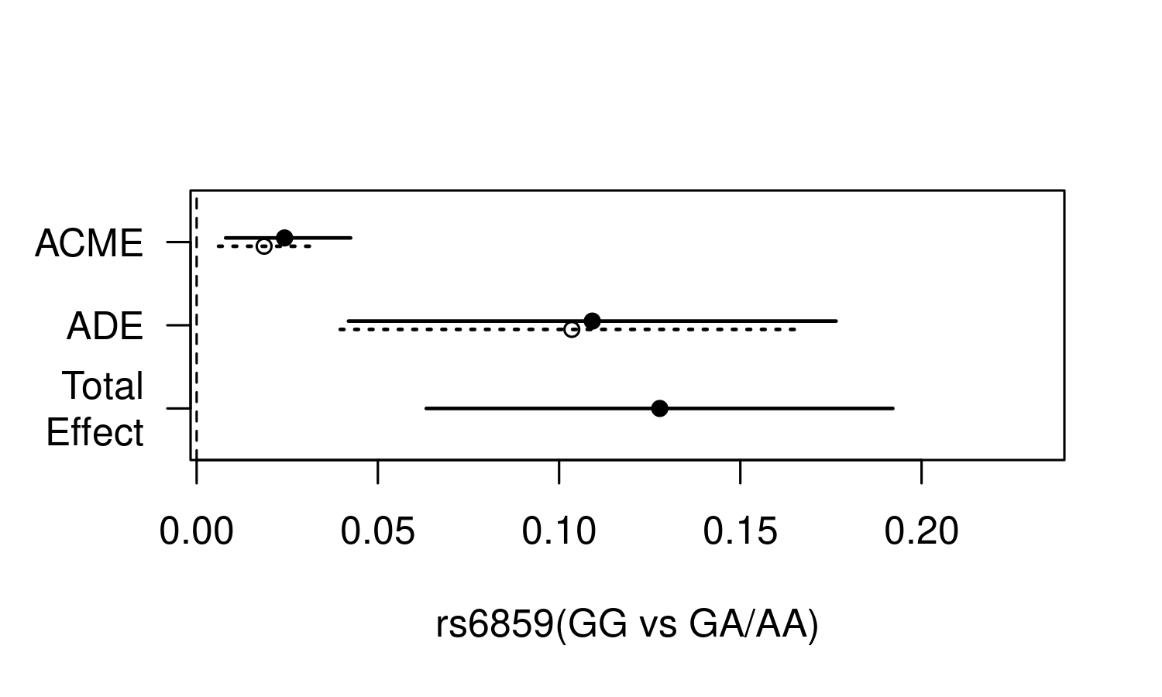

**Supplementary Figure S7.** Contrasts in estimates of causal mediation analysis for SNP rs6859 between carriers of GA/AA genotype and GG genotype with AD mediated through Ptau-181. Note. The solid black line and dashed line represent estimate of the average causal mediation effects (ACME), average direct effects (ADE), and total effect for carriers and non-carriers, respectively. The circle represents point estimates, and whiskers show the corresponding confidence intervals. **ACME (Average Causal Mediation Effect)** indicates the part of the rs6859 effect that is mediated by pTau-181. **ADE (Average Direct Effect)** refers to the part of the effect that is independent of the pTau-181. **Total Effect** includes both the direct and indirect effects of rs6859. **Proportion Mediated** is calculated as the ratio of the ACME to the total effect, representing the proportion of the effect that passes through pTau-181 due to rs6859.

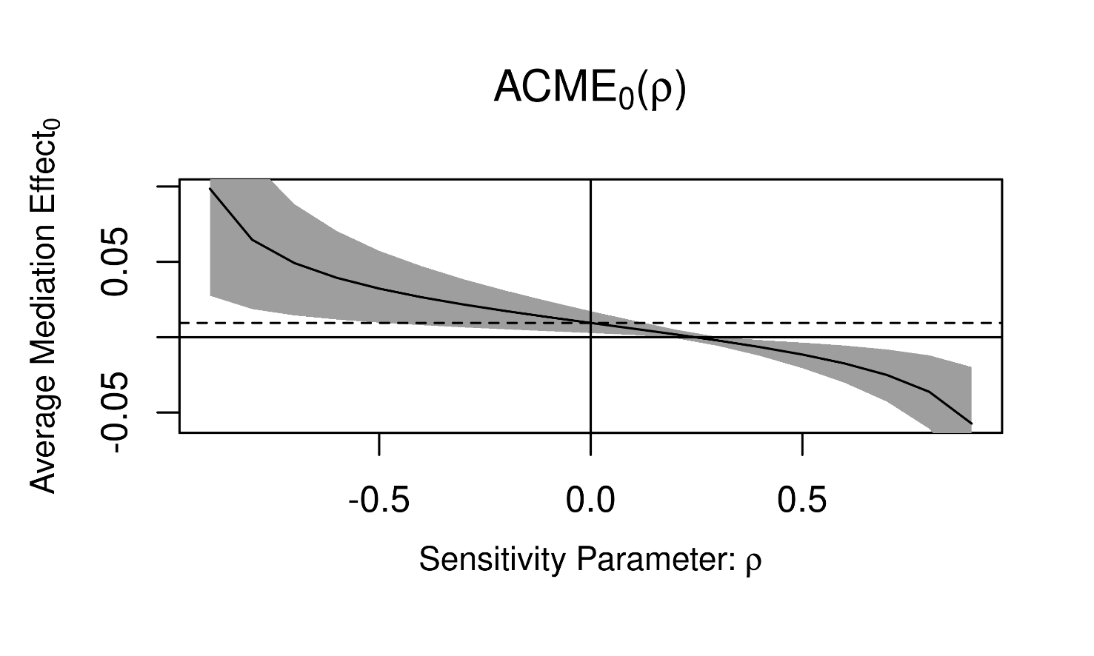

**Supplementary Figure S8.** Sensitivity analysis plot illustrating the model parameter ρ for checking the sequential ignorability assumption. The plot shows the corresponding change in average causal mediation effects (ACME) resulting from changing the sensitivity parameter value (ρ). **ACME (Average Causal Mediation Effect)** indicates the part of the rs6859 effect that is mediated by pTau-181. Note. The solid line and grey area represent the estimate and 95% confidence intervals corresponding to the average causal mediation effects. The dashed line shows the specific ρ values at which the ACME approaches zero.

**Supplementary Table S1.** Estimates from the probit regression model for SNP rs6859 association with AD from the selected AIC model

| **Variable** | **Coefficient** | **95% CI** | **P value** |
| --- | --- | --- | --- |
| rs6859 | 0.25 | 0.11, 0.39 | 0.000*** |
| Education (years) | -0.04 | -0.07, -0.008 | 0.01* |

^*^p<0.05; ^**^p<0.01; ^***^p<0.001.

**Supplementary Table S2.** AIC for all the different models explored for determining associations with the AD and SNP rs6859 using the probit model

| **Model** | **AIC** |
| --- | --- |
| Model 1 (null) | 806.93 |
| Model 2 (full) | 801.10 |
| Model 3 (reduced) | 792.20 |

Note. The null probit model contains only the intercept term. The full probit model contains all the covariates – age, sex, education, race, marriage status, alcohol and smoking history. The reduced model is the best probit model containing rs6859 and education selected using the AIC criterion.

**Supplementary Table S3.** AIC for all the different models explored for determining associations with the standardized Tau and SNP rs6859 using the linear regression model

| **Model** | **AIC** |
| --- | --- |
| Model 1 (null) | 2012.22 |
| Model 2 (full) | 2001.16 |
| Model 2 (reduced) | 1996.78 |

Note. The null linear regression model contains only the intercept term. The full linear regression model contains all the covariates – age, sex, education, race, marriage status, alcohol and smoking history. The reduced model is the best linear regression model containing age, race, rs6859 and sex selected using the AIC criterion.
